# Sleep-like slow waves during resting-state: a promising EEG biomarker of amyloid and neurodegeneration in preclinical Alzheimer’s disease

**DOI:** 10.64898/2026.01.15.26344195

**Authors:** Pierre Champetier, Claudia Albero, Filipa Raposo Pereira, Rubén Herzog, Maximilien Chaumon, Marion Houot, Maxime Locatelli, Aurélie Kas, Marie-Odile Habert, Marc Teichmann, Stéphane Epelbaum, Isabelle Arnulf, Delphine Oudiette, Thomas Andrillon, the INSIGHT-preAD group

## Abstract

**INTRODUCTION:** Growing evidence supports a critical role of sleep slow waves (SW) in Alzheimer’s disease (AD). However, wake SW (sleep-like SW potentially reflecting local intrusions of sleep) remain unexplored in AD.

**METHODS:** 274 older adults with subjective cognitive decline (INSIGHT-preAD cohort, 76.6 ± 3.5 years) underwent i) PET scans for amyloid (A) and neurodegeneration (N), ii) high-density resting-state EEG recordings to detect wake SW, and iii) cognitive assessments. Biomarkers were reassessed two years later. We examined wake SW associations with 1) current A/N status, 2) cognition, and 3) amyloid conversion.

**RESULTS:** A+N−, A−N+ and A+N+ individuals exhibited lower delta wake SW density than A−N− participants. Wake SW amplitude 1) was higher in A+N+ than A−N− individuals, 2) correlated with poorer cognition, and 3) predicted A− to A+ conversion.

**DISCUSSION:** Wake SW represent promising early EEG biomarkers for AD pathology and amyloid conversion, facilitating risk stratification before cognitive decline onset.

## 1. Background

The prevalence of Alzheimer’s disease (AD), the first cause of dementia, continues to rise with the aging of the world population. Current models project over 150 million dementia cases by 2050, compared to 57 million in 2019,^1^ highlighting the escalating societal and public health challenge posed by this condition. At the biological level, AD is characterized by neuropathological changes including amyloid-β deposition (A), pathologic tau accumulation (T), and neurodegeneration (N).^2^ These pathological hallmarks start to accumulate gradually, often decades before the onset of cognitive symptoms.^3^ Clinically, patients exhibit a progressive decline in cognitive abilities, most notably memory, with other domains such as spatial orientation, reasoning, attention, and language also affected.^4,5^ Importantly, AD symptomatology is increasingly viewed as a continuum from subjective cognitive decline (SCD), recognized as a potential prodromal stage, to mild cognitive impairment (MCI) and AD dementia.^6^ Neuroimaging studies have shown that brain regions affected by AD are particularly susceptible to neuropathological changes in individuals with SCD.^7^ However, not all individuals with SCD progress along the AD continuum.^8^ Thus, early identification of individuals at high risk for progression to AD is critical in the perspective of neuroprotective interventions, before irreversible damage occurs.

Beyond cognitive deficits, individuals with AD often complain of sleep disturbances, including prolonged sleep latency, frequent nighttime awakenings, and excessive daytime sleepiness.^9^ Emerging evidence in the past two decades has highlighted a bidirectional relationship between AD and sleep disturbances.^10^ While amyloid and tau accumulation may disrupt subcortical sleep-wake regulation and cortical slow-wave (SW) generation, sleep disruptions are also increasingly recognized as a potential risk factor for AD.^11^ Longitudinal studies have revealed that a range of sleep impairments (e.g., increased sleep latency, fragmentation, reduced sleep efficiency), whether assessed subjectively^12–18^ or objectively,^19–25^ affect both brain integrity and cognitive abilities years or even decades after, increasing the risk of dementia. More specifically, disruption of sleep SW may impair the removal of amyloid deposits,^26^ potentially accelerating AD progression. Accordingly, cognitively unimpaired older adults with low baseline SW activity (SWA) showed faster amyloid accumulation on PET imaging over a 3.7-year follow-up,^23^ although this result was not replicated in another shorter longitudinal study.^27^ Moreover, experimental SWA disruption during sleep has been shown to increase amyloid-β40 levels in cerebrospinal fluid (CSF) the following morning.^28^

Interestingly, sleep-like SW can occur locally during wakefulness as shown by recent evidence in rodents^29^ and humans.^30–34^ These wake SW share similarities with those observed during non-rapid eye movement (NREM) sleep. For instance, they also seem to reflect neuronal silencing in rodents^29^ and humans.^34^ Besides, their occurrence and amplitude increase after sleep deprivation or disruption^30,31^ similarly to sleep SW. However, wake SW differ from sleep SW in being smaller in amplitude and more local in time and space.^36^ This means that wake SW could represent transient sleep-like events confined to specific brain regions, while the rest of the brain exhibits typical wake EEG activity. Importantly, wake SW may affect behavior given their association with errors and attention lapses across different cognitive tasks.^32,34,37^ The concept of wake SW is particularly relevant in the context of AD, as AD patients exhibit 1) daytime sleepiness^38^ and attention deficits,^39^ as well as 2) EEG slowing during wakefulness with increased activity in low-frequencies (delta and theta bands).^40,41^ Despite this, wake SW have never been formally investigated in SCD/AD patients.

In this study, we aimed to fill this gap by investigating wake SW in individuals with SCD from the INSIGHT-preAD cohort. Our primary objective was to explore the relationship between wake SW and two AD pathological hallmarks: A and N. Specifically, we aimed to assess whether wake SW metrics could serve as early predictors of pathological progression. Secondly, we examined if resting-state wake SW features differed between pre- and post-task periods. Finally, we evaluated the associations between wake SW and cognitive performance as measured by neuropsychological tests.

## 2. Methods

### Participants

Participants were enrolled in the monocentric observational INSIGHT-preAD study conducted at the Institute for Memory and Alzheimer’s Disease (IM2A) at Pitié-Salpêtrière University Hospital in Paris, France. The INSIGHT-preAD cohort consists of 318 cognitively unimpaired older adults (aged 70–85 years) with SCD, followed up over a 5-year period. Detailed inclusion and exclusion criteria have been previously described.^42^

To be included, participants had to meet the following criteria: i) presence of memory complaints for at least 6 months, ii) normal global cognitive efficiency (MMSE, Mini-Mental State Examination score ≥ 27 and Clinical Dementia Rating score = 0), and iii) no signs of episodic memory deficits (FCSRT, Free and Cued Selective Reminding Test total recall score ≥ 41). Exclusion criteria included being under guardianship or in a nursing facility, having a diagnosis of prodromal AD or any other neurological disorder, illiteracy, or any contraindication to performing brain MRI.

Comprehensive data were collected at baseline and during regular follow-ups over a 5-year period, including demographic, cognitive, functional, biological, genetic, genomic, imaging (structural and functional MRI, ^18^F-FDG PET, and ^18^F-florbetapir PET), and electrophysiological assessments. Education level was categorized into two groups: individuals with less than 12 years of education were classified as having a low education level, while those with 12 or more years were considered to have a high education level.

The study protocol was approved by the ethics committee of Pitié-Salpêtrière University Hospital (IDRCB: 2012-A01731-42), and all participants provided written informed consent in accordance with the Declaration of Helsinki. Our analyses focused on two time points (baseline and 2 years later) where EEG and imaging data were available. Details on the specific time points used for each analysis are provided below in the Statistical analyses section. After excluding artifact-contaminated EEG recordings (see EEG acquisition section), data from 274 participants were analyzed at baseline, and year-2 follow-up data were available for 247 of them. Demographic data are summarized in **Table 1**.

**Table 1.** Participant characteristics.

|  | Baseline | 2 years later |
| --- | --- | --- |
| **Demographics** |  |  |
| Number of individuals | 274 | 247 |
| Age (years) | 76.6 ± 3.5 | 78.3 ± 3.4 |
| Sex ratio (% of women) | 62.8 | 60.3 |
| Education (% of individuals with high education) | 68.2 | 70.9 |
| ApoE4 genotype (% of ApoE4 carriers) | 19.3 | 20.6 |
| **Neuroimaging data** |  |  |
| A status: nb (%) of A+ individuals * | 78 (28.6) | 82 (34.6) |
| Mean SUVR <sup>18</sup>F-florbetapir | 0.68 ± 0.14 | 0.70 ± 0.14 |
| N status nb (%) of N+ individuals # | 69 (25.3) | 80 (34.2) |
| Mean SUVR <sup>18</sup>F-FDG | 2.12 ± 0.21 | 2.05 ± 0.18 |
| Nb (%) of A-N- individuals | 147 (53.8) | 109 (46.6) |
| Nb (%) of A+N- individuals | 57 (20.9) | 45 (19.2) |
| Nb (%) of A-N+ individuals | 48 (17.6) | 43 (18.4) |
| Nb (%) of A+N+ individuals | 21 (7.7) | 37 (15.8) |
| **Cognition** |  |  |
| MMSE (total score, /30) | 28.6 ± 1.0 | 28.8 ± 1.3 |
| FCSRT (immediate total score, /48) | 46.1 ± 2.0 | 46.6 ± 2.0 |
| FAB (total score, /18) | 16.5 ± 1.6 | 16.6 ± 1.6 |

Data are presented as mean ± standard deviation for continuous metrics. Education level was categorized into two groups: individuals with less than 12 years of education were classified as having a low education level, while those with 12 or more years were considered to have a high education level. \*: <sup>18</sup>F-florbetapir-PET data were not available for one participant at baseline, ten participants at the year-2 follow-up. #: <sup>18</sup>F-FDG PET data were not available for one participant at baseline, and for thirteen participants two years later. Abbreviations: A (Amyloid), FAB (Frontal Assessment Battery at Bedside), FCSRT (Free and Cued Selective Reminding Test), FDG (FluoroDeoxyGlucose), MMSE (Mini-Mental State Examination), N (Neurodegeneration), SUVR (Standardized Uptake Value ratio).

### Cognitive composite scores

All neuropsychological assessments were performed in the morning. To reduce the number of cognitive metrics, cognitive composite scores were computed by standardizing outcome measures for each neuropsychological test on each assessment using baseline mean and standard deviation for the entire group. We focused our analyses on three composite scores evaluating three key cognitive domains: memory, attention, and executive functions.

1. The **memory** composite score was computed by averaging the standardized and normalized scores on the FCSRT (immediate and delayed free recalls), Rey Complex Figure Test (3min and 30min scores), Delayed Matching to Sample 48 test (DMS48, immediate and 1h delayed score), Memory Binding Test (MBT, immediate and delayed scores for list A and B combined).
2. The **attention** composite score was computed by averaging the standardized and normalized scores on the Trail Making Test (TMT, total error at part A) and the Digital Span Direct (numeric and visuo-spatial).
3. The **executive functions** composite score was computed by averaging the standardized and normalized scores on the TMT (number of perseverative errors at part B, and time difference between parts B and A), Frontal Assessment Battery (FAB, total score), and lexical verbal fluency (for the letter “p”).

### Neuroimaging exams

Baseline and 2-year follow-up PET scans (Gemini XLS, Philips) included amyloid imaging with 370 MBq ^18^F-florbetapir (50 minutes post-injection, 20-min acquisition) and FDG imaging as a proxy for neurodegeneration with 2 MBq/kg ^18^F-FDG (30 minutes post-injection, 15-min acquisition). Reconstructed images were analyzed using semi-automatized pipelines developed by the Centre d’Acquisition et Traitement des Images (CATI) (http://cati-neuroimaging.com) with BrainVISA (http://brainvisa.info/web/index.html).

Two pipelines were developed: one at each time point for FDG acquisition (as described in^42,43^) and one longitudinal for amyloid data. Each pipeline includes the following five steps: 1) segmentation of the anatomical MRI, 2) application of partial volume effect correction to the PET images, 3) spatial normalization in the chosen space, 4) intensity normalization, and 5) extraction of standardized uptake value ratio (SUVr). Thus, each 3D T1-weighted MRI was segmented using SPM12 to obtain masks for gray matter, white matter, CSF, skull, and scalp. These images were then spatially normalized to the Tissue Probability Map (TPM) template, aligning them to MNI space. After a partial volume effect correction applied in PET space using RBV-sGTM method,^44^ PET images were rigidly registered to their corresponding T1 MRI and subsequently transformed into MNI space using the transformation fields obtained during MRI normalization.

In the pipeline for ^18^F-FDG data, the reference region comprised the white matter in the pons of the subject. In the pipeline for amyloid data, an ad hoc reference region suited for longitudinal studies was defined for each subject, combining the pons, the entire cerebellum, and the supratentorial white matter (as described in^45^). Intensity normalization was performed by dividing each voxel’s activity by the mean activity extracted from the reference region of the selected pipeline. Finally, regional SUVr values were extracted using AAL atlas.^46^ Amyloid SUVrs were obtained using a CATI-specific atlas that includes bilateral precuneus, cingulum (posterior and anterior), and parietal, temporal, and orbito-frontal cortices ^43^. The mean SUVr was calculated by averaging the mean activity of the 12 cortical regions of interest defined in the CATI atlas. Cortical metabolic indices were also calculated in four bilateral regions of interest (ROI) specifically affected by AD:^47^ posterior cingulate cortex, inferior parietal lobule, precuneus, and inferior temporal gyrus, using the pons as the reference region. All data and processing steps were checked by the CATI engineers. Individuals with ^18^F-florbetapir PET SUVr > 0.6823 were classified as amyloid-positive (A+), and those with ^18^F-FDG PET SUVr < 1.9763 as neurodegeneration-positive (N+). These thresholds were obtained by adapting those from Habert et al.^43^ and Gaubert et al.^48^ to our processing pipeline using linear regression.

### EEG acquisition

This study focused on EEG and other neuroimaging data collected at baseline and 2 years later. Recordings were made during the afternoon, using a high-density 256-channel EGI system (Electrical Geodesics Inc.), with a sampling rate of 250 Hz and a vertex reference. At each time point, EEG data acquisition included two 2-min resting-state sessions, separated by a 20-min memory task (old/new recognition paradigm using FCSRT words performed 4 to 6 hours earlier as old items). During the two resting-state sessions (i.e., before and after the task), participants were instructed to stay awake and relaxed while alternating between 30-s of eyes closed and eyes open segments. EEG preprocessing was made using a pipeline that automatically removed artifacts and extracted EEG measures. All non-scalp electrodes were excluded from analysis, restricting the dataset to the 192 scalp channels. A band-pass filter (0.1–40 Hz) and a notch filter (50 Hz, 100 Hz and 150 Hz) were applied. Blinks were identified using ft_artifact_eog function of FieldTrip. Data were segmented into 10-s epochs to identify bad channels. A channel was marked as bad if at least 25% of its epochs showed a z-score > 4 (relative to other channels) for any of the following voltage features: standard deviation, maximum absolute value, or kurtosis. This procedure was applied separately to frontal electrodes and to the rest of the scalp, using blink-free periods. Bad channels were interpolated. ICA was then performed after excluding 1-s artifacted segments and high-pass filtering at 1-Hz. Eye independent components (IC) were automatically labeled and removed using SASICA. A bespoke procedure was applied to identify and remove cardiac IC in the absence of ECG channel (https://github.com/PierreChampetier). In short, cardiac events (PQRST) were detected across all ICs. Cardiac ICs were identified based on i) the distribution of multiple features (e.g., skewness, kurtosis) extracted from detected cardiac events (e.g., RR intervals, R-wave amplitude), and ii) sanity checks such as physiological heart rate derived from R-peak counts. To evaluate ICA quality, blinks were also automatically identified in preprocessed data using the same threshold, and a ratio between the number of blinks before and after ICA was computed. Recordings with i) density of blinks after ICA > 2/min, and a ratio before/after ICA < 2, or ii) more than 15% of samples > 150 µV were excluded (12% of the recordings).

### Wake SW detection

Wake SW were detected using a similar procedure to that in Andrillon et al.^32^, based on previously devised algorithms for automatic detection of SW during NREM sleep.^49^ The preprocessed EEG signal was first re-referenced to the average of the left and right mastoid electrodes (E94 and E190), in accordance with established sleep recording guidelines.^50^ The signal was then down-sampled to 128 Hz and band-pass filtered using a Butterworth filter in the passband (1–10 Hz). Negative peaks in the filtered signal were used to detect all waves, and several parameters were extracted for each wave: start and end points (defined as zero-crossings before the negative peak and after the positive peak), negative and positive peak amplitudes and their time positions, peak-to-peak amplitude, downward and upward slopes, and frequency (1/duration). To minimize false detections caused by artifacts such as potential remaining blinks (which often exhibit a prominent positive component), waves with a positive peak exceeding 75 μV or occurring within 1 s of large-amplitude events (>150 μV absolute amplitude) were excluded. Only waves with a peak-to-peak amplitude above 5 µV were kept. Waves with a frequency (1/duration) between 1 and 4 Hz were identified as delta SW, and those with a frequency between 4 and 7 Hz were identified as theta SW. Finally, channels with a SW density below 5/min were removed from analyses (< 1% of EEG channels). For both delta and theta wake SW, our metrics of interest were their density and amplitude.

### Statistical analyses

All statistical analyses were performed using Matlab R2023a. Linear (LM) and linear mixed effect models (LME) were performed on each EEG channel, after controlling for age, sex and education level. A cluster-based permutation method was used to control for multiple comparisons (see below). Such an approach was used in the five following analyses:

1. The effect of the A/N status on wake SW features at rest (before the task) was tested by comparing the A+N−, A−N+ and A+N+ groups to the control group A−N−. Analyses were conducted at the year-2 follow-up to maximize statistical power, as the smaller group (A+N+) had larger sample sizes at this time point (**Table 1**).
2. The modulation of wake SW features by the cognitive task was assessed by comparing the two resting-state recordings (pre- and post-task). To investigate potential effects of A/N status, additional analyses were performed separately within each group. Thus, these analyses were also performed at year-2 follow-up. For each EEG channel, LME models were used to account for within-subject effects.
3. The relationships between wake SW features at rest (before the task) and cognitive functioning (at baseline) were also assessed. To do so, the three cognitive composite scores (memory, attention, executive functions) were added to a linear model performed on each EEG channel, and the main effect of each composite score was extracted. Analyses were performed at baseline to maximize the sample size.
4. For exploratory purposes, we also examined the impact of demographic variables (age, sex and educational level) on wake SW features at rest (before the task) by analyzing the main effect of each of these variables in linear models without additional variables. Similarly, analyses were performed at baseline to maximize the sample size.
5. To assess the predictive value of pre-task wake SW features at baseline for longitudinal amyloid changes, participants were classified based on their A status evolution between baseline and the year-2 follow-up. Using linear models, individuals that were A− at baseline and became A+ two years later (A−_A+ group) were compared to those who remained A−(A−_A− group) and to those who remained A+ (A+_A+ group). Four individuals that were A+ at baseline and became A− two years later were removed from these analyses, as this change was most likely due to measurement variability in PET imaging near the amyloid positivity threshold.

For each analysis, we provided a quantified visual representation of the modulations of wake SW features by the variable of interest. When comparing two groups (e.g., A+N− vs A−N−), we plotted topographies of the differences between the mean values of wake SW metric of the two groups. When testing the effect of a continuous metric (e.g., age) on wake SW features, we plotted topographies of the t-values of the variable of interest. Channels belonging to a significant cluster were identified with a cluster-based permutation approach, and are highlighted on each topoplot. The cluster-based permutation approach was derived from Maris & Oostenveld.^51^ For each analysis, the t and p-values were calculated for all EEG channels separately. Candidate clusters were defined as at least two neighbouring electrodes with p-value below 0.025 (cluster alpha). For each candidate cluster, the t-values of all electrodes within the cluster were summed to compute a cluster statistic. Data of the metric of interest (e.g., the A/N status) were then shuffled to create 1000 permuted datasets. For each permuted dataset, a similar procedure was applied, and the candidate cluster with the maximal absolute cluster statistics was identified to form a null distribution. The cluster statistics of candidate clusters from the real dataset were compared to the null distribution, and a Monte Carlo p-value was derived from this comparison. A p_cluster_ < 0.05 indicates that the cluster statistic lies in the extreme 5% of the permutation distribution (i.e., below the 5th percentile for negative clusters or above the 95th percentile for positive clusters).

Finally, we assessed whether wake SW amplitude at baseline could predict which A− individuals would convert to A+ during the longitudinal follow-up. Since EEG recordings collected in clinical practice typically involve few EEG channels, we used the averaged wake SW amplitude across the whole scalp to avoid focusing on a specific scalp region. Data were split into training (80%) and testing (20%) sets, maintaining the proportion of A-_A+ in both subsets. In the training set, conversion from A− to A+ was predicted using logistic regressions with either: 1) baseline averaged amplitude of wake SW (delta or theta) alone, 2) baseline ^18^F-florbetapir PET SUVr alone, or 3) a combined model including both PET SUVr and the averaged amplitude of delta and/or theta wake SW. An optimal threshold was derived from the training set using the Youden index (maximizing sensitivity and specificity) and applied to the testing set to compute sensitivity, specificity, and balanced accuracy (accounting for class imbalance). This procedure was repeated using 100 iterations of 5-fold cross-validation to obtain mean values and 95% confidence intervals for each metric.

## 3. Results

### Wake SW features are modulated by A and N status

We first evaluated how wake SW differ depending on the A/N status. To enable more reliable group comparisons, analyses used year-2 follow-up data, which showed greater distributional balance than baseline. Characteristics of each A/N group (A−N−: n=109, A+N−: n=45, A−N+: n=43, A+N+: n=37) are described in **Supplementary Table 1**. We quantified wake SW at rest, before the cognitive task (see **Fig. S1A** for topographies of wake SW density and amplitude in the delta and theta bands). First, we compared wake SW characteristics in individuals positive for amyloid and/or neurodegeneration (A+N−, A−N+, and A+N+) to those negative for both biomarkers (A−N−, **Fig. 1**). Compared to the A−N− group, all biomarker-positive groups showed reduced delta wake SW density, with a widespread cluster covering most of the scalp (A+N−: p_cluster_=0.009; A−N+: p_cluster_<0.001; A+N+: p_cluster_=0.005). The A+N+ group also exhibited greater amplitude of wake SW compared to A−N− individuals, in both the delta (p_cluster_= 0.023) and theta (p_cluster_=0.015) bands, with clusters peaking in the centro-parietal region. A−N+ and A+N− groups did not differed in averaged wake SW amplitude compared to A−N− group, but the distribution of wake SW amplitude in A−N+ individuals exhibited an intermediate pattern between A−N− and A+N+ groups (**Fig. S2**).

**Figure 1:**
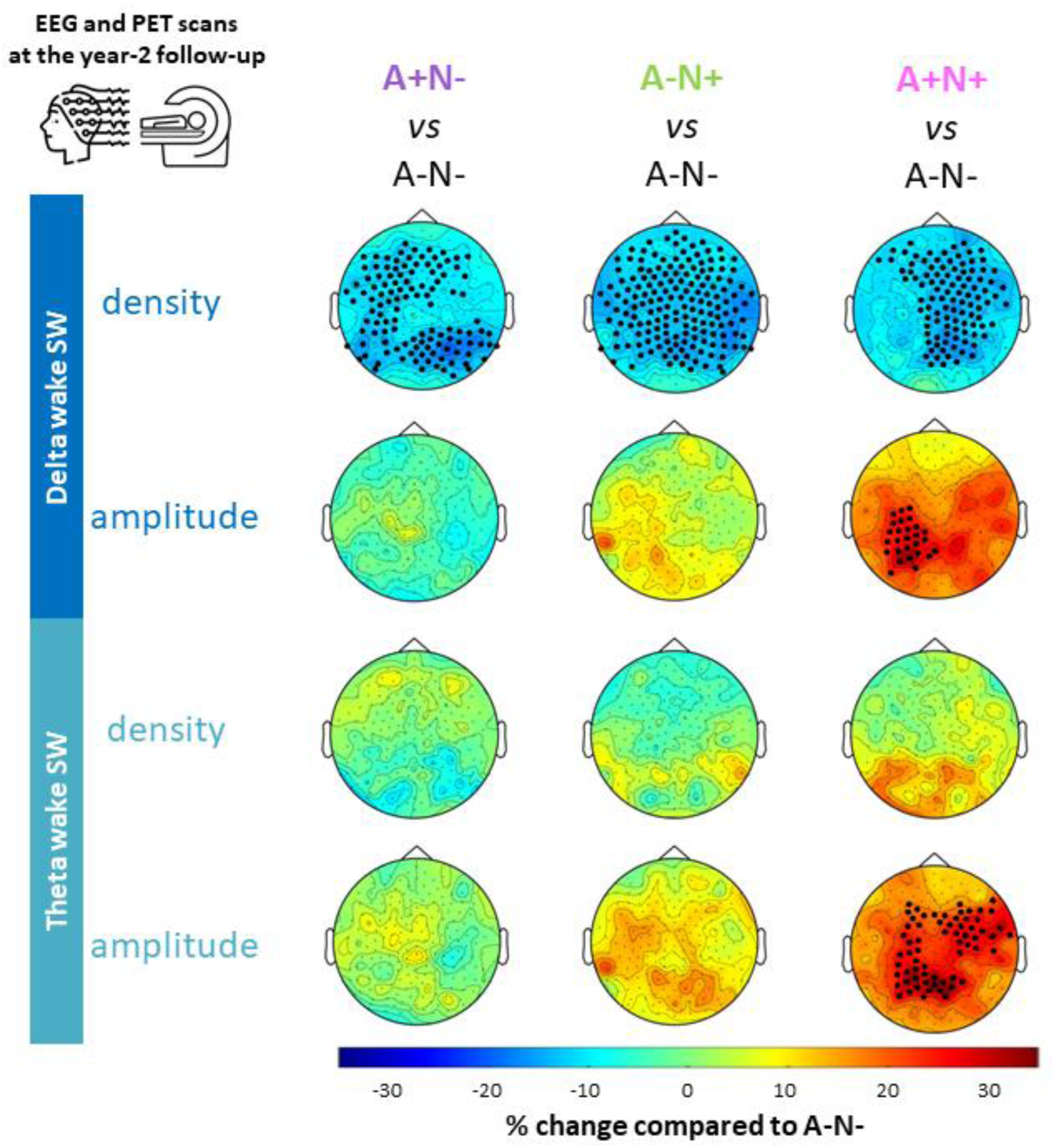
Effect of A and N status on resting-state wake SW features. Topographies representing the proportional change in mean resting-state wake SW metrics (density and amplitude) for each test group (A+N−, A−N+ and A+N+) relative to A−N− individuals (e.g., 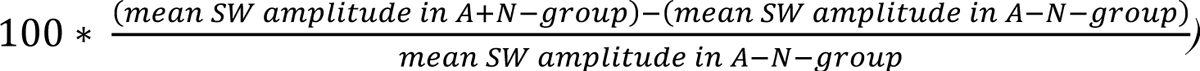. indicate higher density (or amplitude) in the test group compared to A−N− individuals, while negative values denote lower density (or amplitude). Black dots (•) represent significant clusters of electrodes using a cluster-based permutation approach after controlling for age, sex and education level using linear models. Data at the year-2 follow-up were used as group distributions were more balanced compared to baseline (**Table 1**). <u>Abbreviations</u>: A (Amyloid), N (Neurodegeneration), SW (Slow Waves).

### Wake SW features are modulated following a cognitive task

We next assessed whether engaging in a cognitive task modulated wake SW features at rest (before/after comparison). We first compared the two resting-state recordings (pre- and post-task) across all participants, independently of the A/N status. Topographies of wake SW features in the post-task session are represented in **Fig. S1B**. All features were strongly affected by the task (**Fig. 2**). Following the task, participants had a lower density of delta wake SW and a higher wake SW amplitude in both delta and theta bands over the whole scalp (all p_cluster_<0.001) compared to the pre-task recording. They also exhibited greater density of theta wake SW, predominantly in the posterior region (p_cluster_<0.001). Similar task-related changes were observed when the analysis was performed separately within each A/N group, except that A+N+ individuals did not show a significant increase in the amplitude of delta wake SW between the two resting-state recordings (**Fig. S3**).

**Figure 2:**
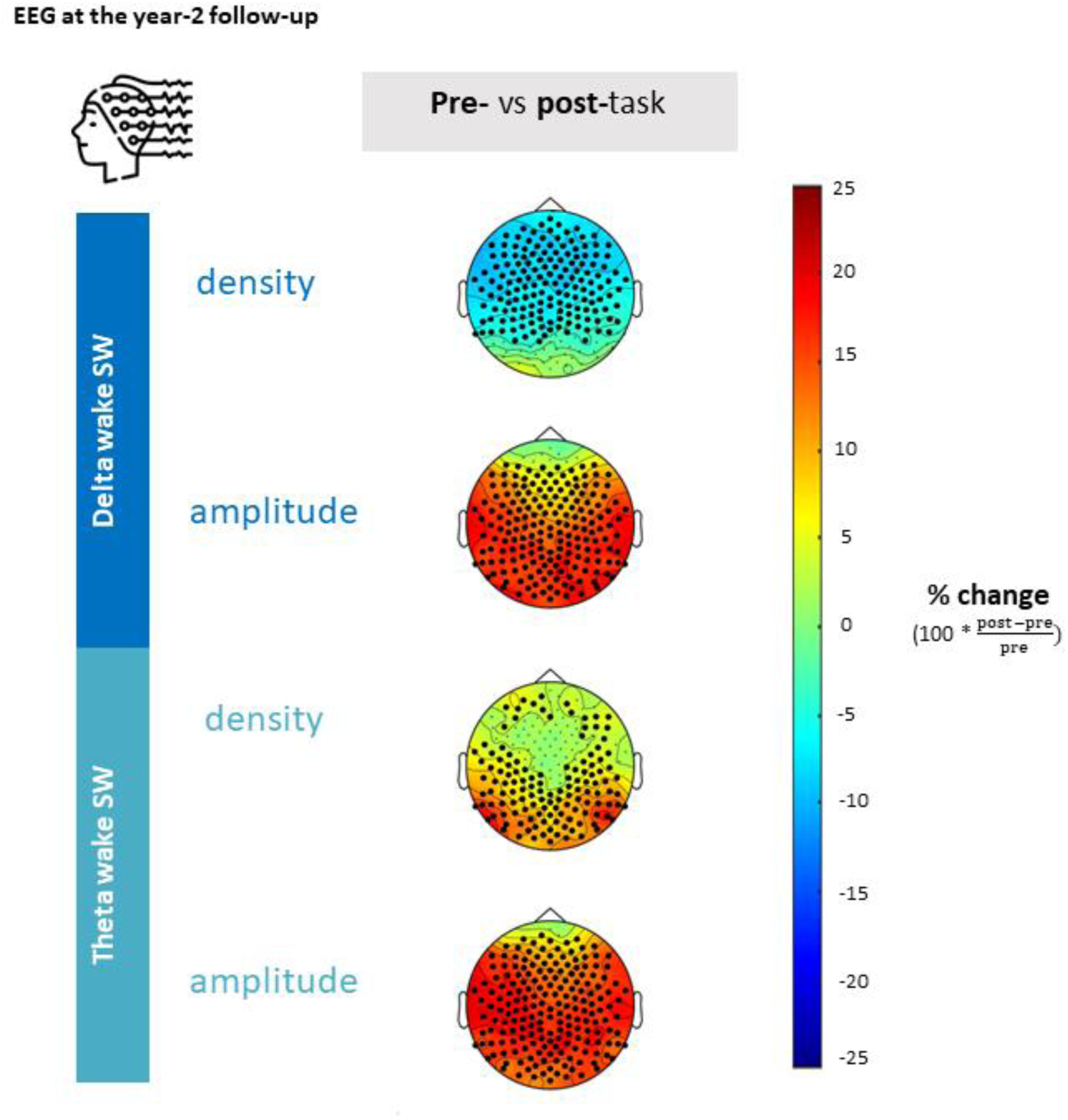
Changes in wake SW features between the pre- and post-task resting-state recordings. Topographies representing the proportional change in mean wake SW metrics (density and amplitude) between the two resting-state recordings. Positive values indicate an increase in the post-task relative to the pre-task resting-state, while negative values denote a decrease. Black dots (•) represent significant clusters of electrodes using a cluster-based permutation approach after controlling for age, sex and education level using linear mixed models. <u>Abbreviation</u>: SW (Slow Waves).

### Higher amplitude of wake SW is associated with poorer memory at neuropsychological assessment

We then investigated the associations between wake SW features at rest and neuropsychological assessment scores using baseline data. Overall, wake SW density was not related to cognition, but higher amplitude of wake SW (in both delta and theta bands) was associated with poorer cognition (**Fig. 3**). The strongest effect was found for the memory composite score, with large significant clusters over the frontal region extending to the central region for the delta band (delta: p_cluster_=0.004, theta: p_cluster_=0.010). Smaller significant clusters were found in the frontal and occipital regions for attention (delta: p_cluster1_=p_cluster2_=0.028, theta: p_cluster_=0.020), and executive functions (delta: p_cluster_=0.021, theta: p_cluster_=0.045).

**Figure 3:**
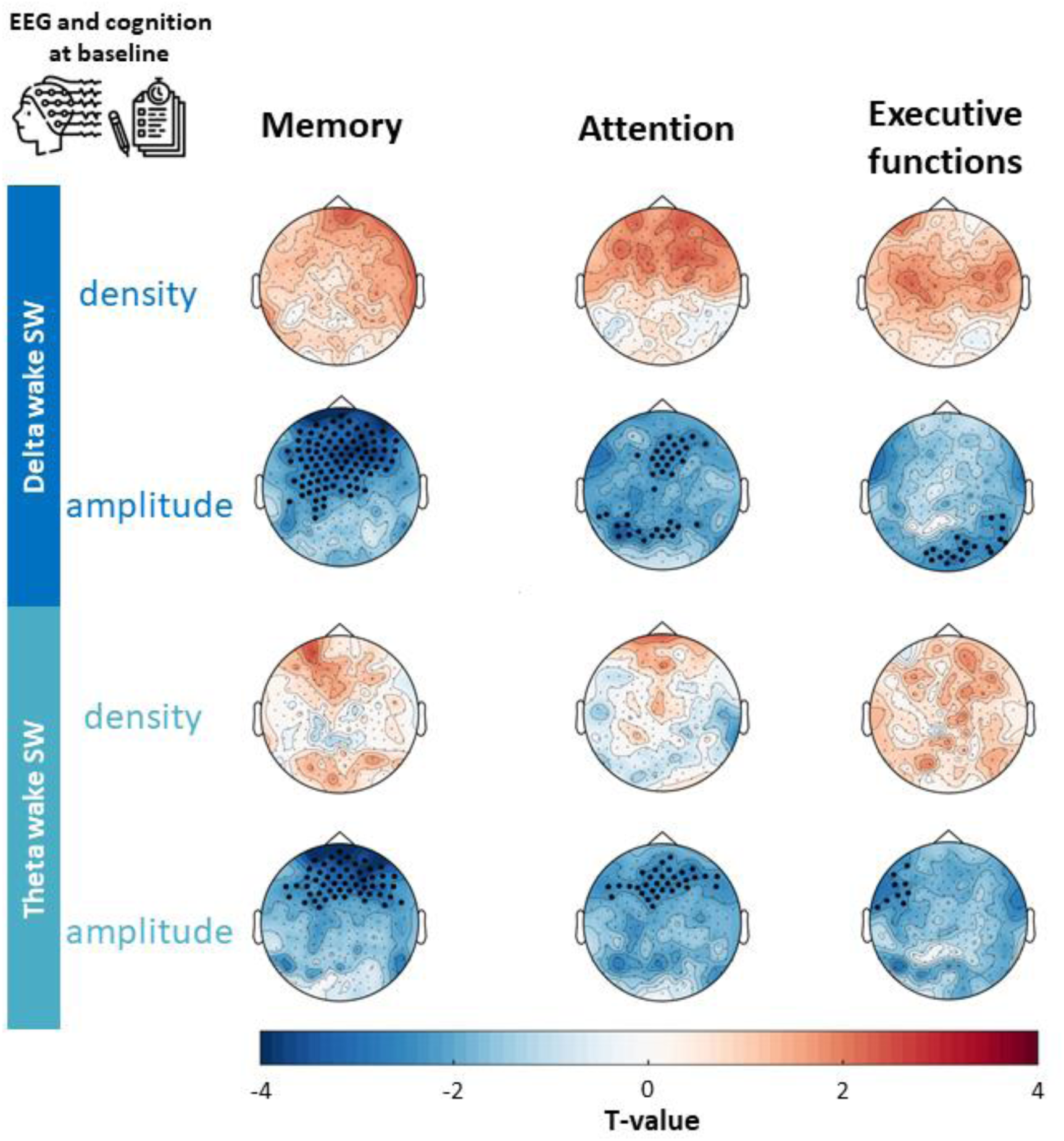
Associations between resting-state wake SW features and neuropsychological assessment scores at baseline. Statistical maps (t-values) of the associations between wake SW features at rest and three cognitive composite scores (memory, attention and executive functions). Red indicates positive associations, while blue denotes negative associations. Black dots (•) represent significant clusters of electrodes using a cluster-based permutation approach after controlling for age, sex and education level using linear models. <u>Abbreviation</u>: SW (Slow Waves).

### Wake SW features differ between men and women

We conducted exploratory analyses on baseline data to test the potential effect of sex, education, and age on wake SW features at rest. We found strong sex-differences in wake SW at rest (**Fig. 4**). Compared to women, men had more theta wake SW in the posterior part of the scalp (p_cluster_ < 0.001). Men also exhibited delta and theta wake SW with higher amplitude across the whole scalp (expect in the prefrontal region), with increases reaching up to 64% in the centro-parietal region (p_cluster_ < 0.001 for both delta and theta bands). A minor effect of educational level was also found, with individuals with higher education having a 5% lower theta wake SW density in one small clusters (p_cluster1_ = 0.012). No significant effect of age was found.

**Figure 4:**
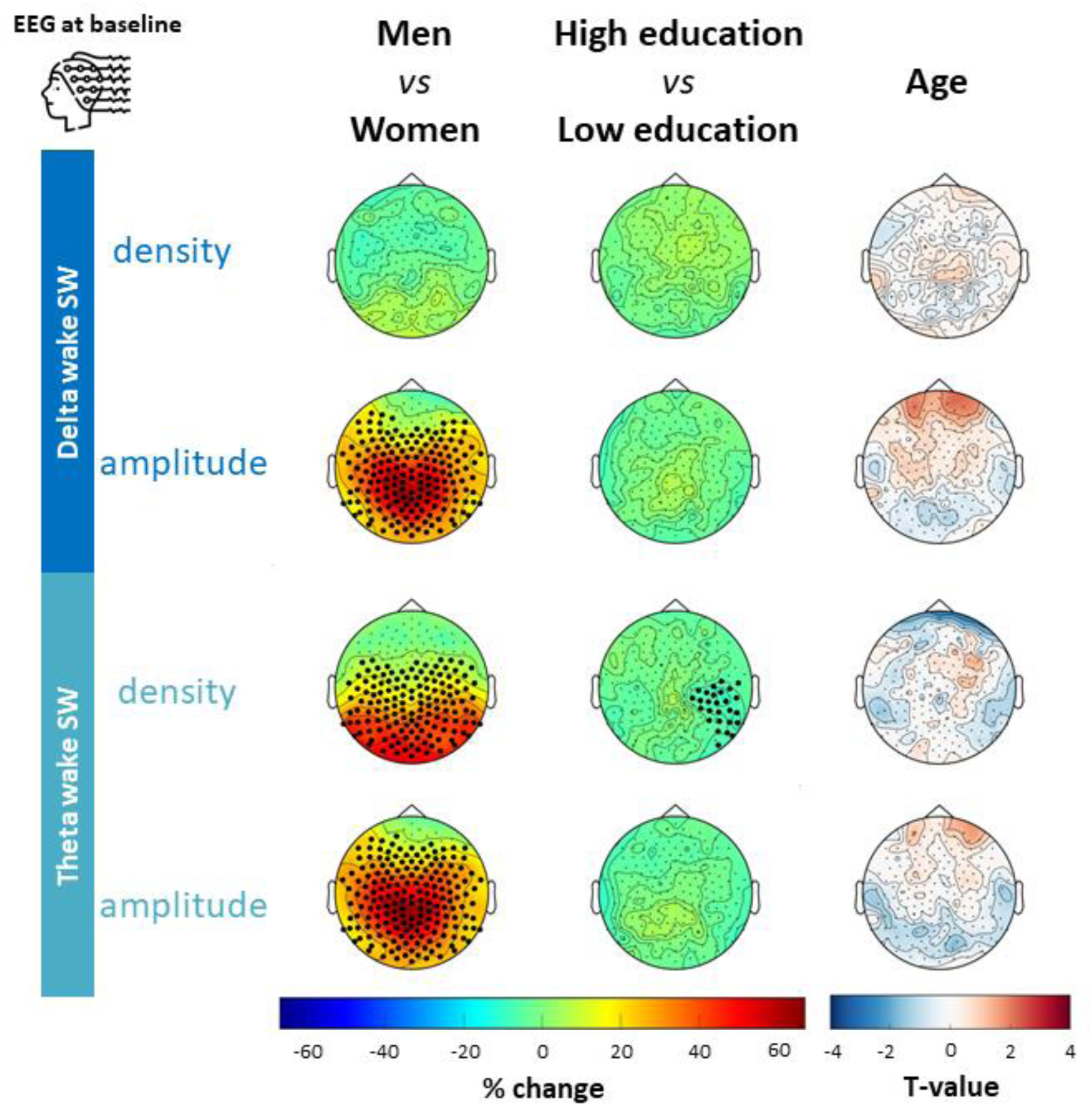
Effect of demographic variables on resting-state wake SW features at baseline. (Left): Topographies representing the proportional change in mean resting-state wake SW metrics (density and amplitude) in men relative to women, and in individuals with high education levels compared to those with low education levels (e.g., 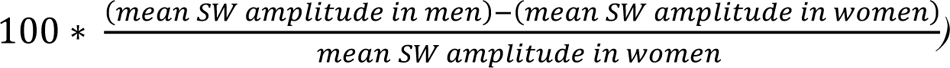. higher density (or amplitude) in men and individuals with higher education levels relative to women and individuals with lower education levels, while negative values denote lower density (or amplitude).(Right): Statistical maps (t-values) of the associations between wake SW features at rest and age. Red indicates positive associations, while blue denotes negative associations. For all topoplots, black dots (•) represent significant clusters of electrodes using a cluster-based permutation approach. <u>Abbreviations</u>: SW (Slow Waves).

### Amplitude of wake SW predicts the conversion from A− to A+ within two years

We next assessed the potential of resting-state wake SW metrics as early indicators of pathological progression. To this end, we stratified participants according to changes in A status between baseline and the 2-year follow-up, yielding three groups: stable A− (A−_A−, n=157), converters (A−_A+, n=16), and stable A+ (A+_A+, n=63). Characteristics of the three groups are described in **Supplementary Table 2**.

Although both groups were amyloid-negative at baseline, the A−_A+ group had higher wake SW amplitude at baseline in both delta (p_cluster_=0.004) and theta (p_cluser_<0.001) bands compared to A−_A− participants (**Fig. 5**). This difference spanned across most EEG channels (especially for the theta band) with increases up to 55% within a centro-parietal cluster. Similar albeit smaller differences were also found when comparing A−_A+ and A+_A+ groups at baseline in both the delta (p_cluster_= 0.049) and theta (p_cluster_=0.042) bands. Thus, individuals who converted to A+ status over the following 2 years exhibited the highest baseline wake SW amplitude, despite being A− at that time (**Fig. S4**). However, no significant changes in SW density were observed (**Fig. 5**).

**Figure 5:**
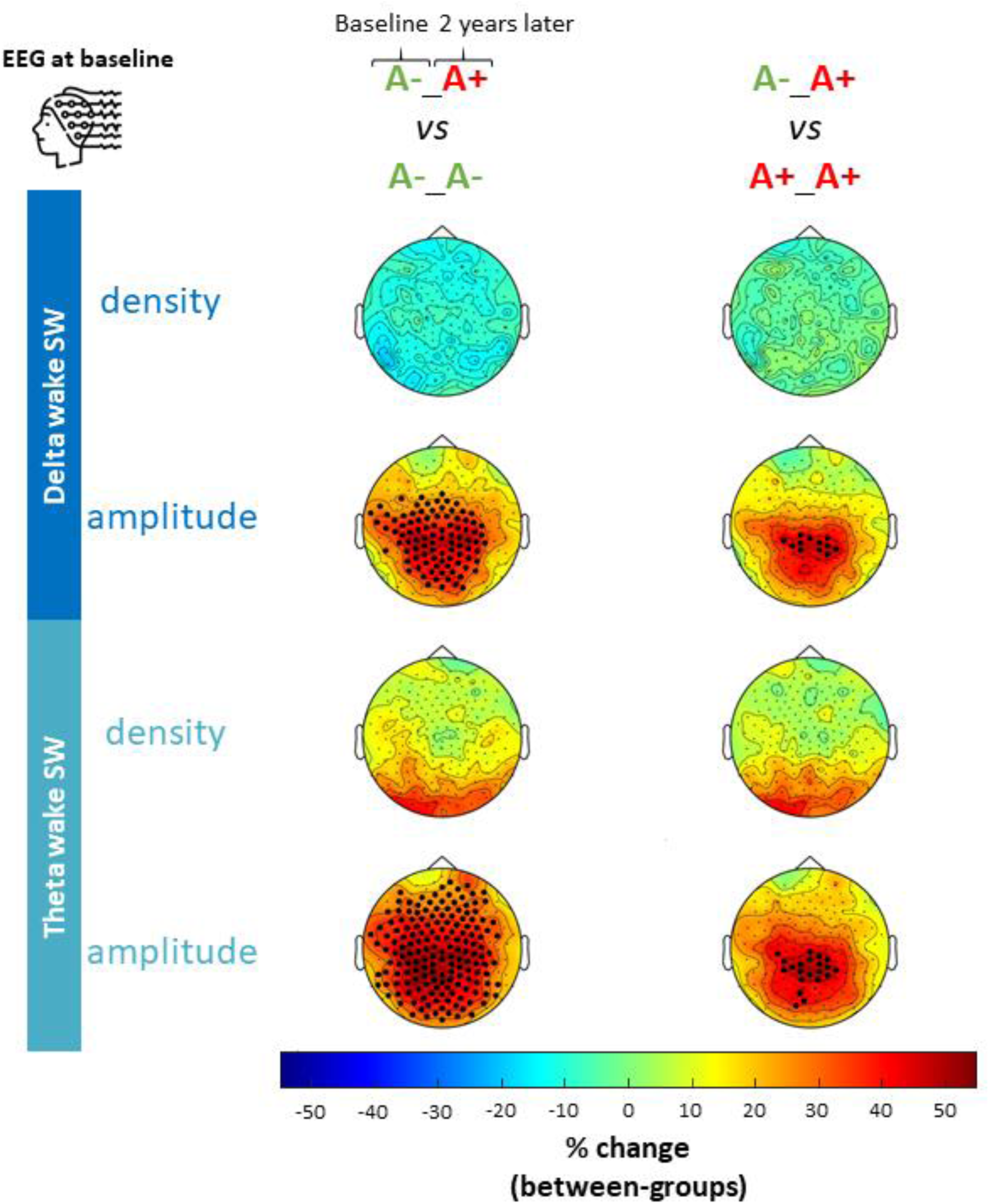
Effect of ongoing amyloid dynamic on resting-state wake SW features quantified at baseline. Topographies representing the proportional change in mean resting-state wake SW metrics (density and amplitude, quantified at baseline) in individuals who converted to amyloid positivity over the 2-year follow-up (A−_A+ group) compared to those who remained either amyloid negative (A−_A− group) or positive (A+_A+ group) (e.g., 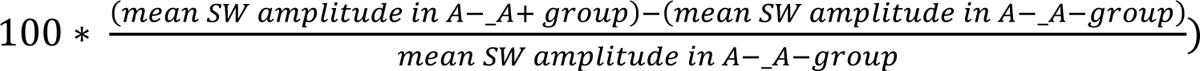. indicate higher density (or amplitude) in the A−_A+ group, while negative values denote lower density (or amplitude). Black dots (•) represent significant clusters of electrodes using a cluster-based permutation approach after controlling for age, sex and education level using linear models. <u>Abbreviations</u>: A (Amyloid), SW (Slow Waves).

Longitudinal analyses showed that delta and theta wake SW amplitude increased during the follow-up period in both A−_A− (all p_cluster_<0.001) and A+_A+ (delta: p_cluster_=0.019, theta: p_cluster_=0.020) groups, but remained stable in the A−_A+ group (**Fig. S5**). Consequently, wake SW features in A−_A+ individuals no longer differed from the other groups at the follow-up (**Fig. S6**).

We then evaluated whether baseline wake SW amplitude could predict which A− individuals would convert to A+ using logistic regression with 100 iterations of 5-fold cross-validation. Both delta and theta wake SW amplitude yielded good predictive performance (delta: balanced accuracy=0.71, sensitivity=0.61, specificity=0.81; theta: balanced accuracy=0.73, sensitivity=0.62, specificity=0.85, **Table 2**). Both showed higher specificity but lower sensitivity compared to baseline ^18^F-florbetapir PET SUVr (balanced accuracy=0.81, sensitivity=0.92, specificity=0.69, **Table 2**). Combining PET SUVr and wake SW metrics yielded models with better balance between sensitivity and specificity (PET + delta wake SW: balanced accuracy=0.82, sensitivity=0.85, specificity=0.78; PET + theta wake SW: balanced accuracy=0.82, sensitivity=0.84, specificity=0.80, **Table 2**). However, adding both wake SW features did not improve the model (balanced accuracy of 0.78). Results remained similar when the models included sex, the only covariate we found to affect wake SW amplitude (**Supplementary Table 3**).

**Table 2.** Prediction of individuals who will convert from A− to A+ during the 2-year follow-up period.

|  | **Sensitivity**<br>**(TP rate)** | **Specificity**<br>**(TN rate)** | **Balanced accuracy** |
| --- | --- | --- | --- |
| Delta wake SW amplitude alone | 0.61 [0.50-0.65] | 0.81 [0.81-0.82] | 0.71 [0.66-0.73] |
| Theta wake SW amplitude alone | 0.62 [0.53-0.65] | 0.85 [0.85-0.85] | 0.73 [0.69-0.75] |
| Baseline <sup>18</sup>F-florbetapir PET SUVR alone | 0.92 [0.80-0.95] | 0.69 [0.69-0.70] | 0.81 [0.75-0.82] |
| Baseline <sup>18</sup>F-florbetapir PET SUVR +<br>delta wake SW amplitude | 0.85 [0.73-0.93] | 0.78 [0.75-0.81] | 0.82 [0.75-0.85] |
| Baseline <sup>18</sup>F-florbetapir PET SUVR +<br>theta wake SW amplitude | 0.84 [0.67-0.90] | 0.80 [0.77-0.83] | 0.82 [0.75-0.85] |
| Baseline <sup>18</sup>F-florbetapir PET SUVR +<br>delta wake SW amplitude +<br>theta wake SW amplitude | 0.75 [0.60-0.88] | 0.80 [0.74-0.86] | 0.78 [0.71-0.83] |

*Conversion from A– to A+ status was modeled using logistic regression with baseline predictors (<sup>18</sup>F-florbetapir PET SUVR and/or wake SW amplitude in delta/theta bands), using 100 iterations of 5-fold cross-validation. Mean values and 95% confidence intervals (in brackets) are reported for sensitivity, specificity and balanced accuracy (accounting for class imbalance). Abbreviations: TN (True Negative), TP (True Positive), SUVR (Standardized Uptake Value ratio).*

## 4. Discussion

We investigated sleep-like SW in resting-state EEG of older adults with SCD to understand their roles in early neuropathological processes. We found that wake SW features vary with the presence of two AD pathological hallmarks: A and N. These changes resembled those that occur following a cognitive task. Moreover, higher wake SW amplitude was associated with poorer cognition and predicted conversion to A+ over a 2-year follow-up. Finally, exploratory analyses revealed sex-differences in wake SW features.

### Wake SW modifications are related to A/N status and mirror those following cognitive task

Our cross-sectional analyses revealed that wake SW features are modulated by A and N status. Compared to A−N− participants, individuals with two positive AD biomarkers (A+N+) exhibited the highest wake SW alterations, including reduced density (in delta band) and increased amplitude (in delta and theta bands). Those positive with only one biomarker (A+N− and A−N+) solely showed a lower density of delta wake SW. Consistently, a previous study analyzing the same cohort with more traditional spectral power analyses reported lower delta power in A−N+ individuals compared to A−N− ones.^48^ However, no differences were found in delta or theta power when comparing A−N− group to the other groups.^48^ Other SCD studies focusing on A status showed either higher delta^52^ or theta power^53^ in A+ compared to A− individuals. By focusing on specific events (i.e., wake SW), our approach examines density and amplitude modulations of wake SW separately, whereas spectral analyses yield a single power value per frequency band. Since we found opposite modulations of delta SW density and amplitude, this could explain the lack of spectral power differences reported in some of these studies.

We also observed modulations of wake SW when comparing resting-state activity before and after a memory task (higher delta and theta amplitudes, increased theta density, decreased delta density). Previous studies focusing on the largest wake SW events have reported increases in density and/or amplitude during a task,^30,32^ largely consistent with our findings, except for delta density. This discrepancy may stem from methodological differences, as we included all wake SW exceeding 5 µV. Interestingly, pre-vs post-task wake SW differences mirrored those observed when comparing A+N+ and A−N− groups. Since task-related modulations of wake SW have been associated with sleepiness and mental fatigue,^32^ this resemblance may suggest that A+N+ individuals may display a “fatigue-like” wake SW pattern at rest. Besides, compared to the other groups, A+N+ individuals showed less pronounced changes in wake SW features following the task (except for delta wake SW density), possibly because they already exhibited a ‘fatigue-like’ pattern prior to the task. This could reflect a form of chronic fatigue and increased sleep pressure in this population, consistent with the sleep disruptions and increased daytime sleepiness often found in AD.^9^ Other studies investigating wake SW in the AD continuum are needed to confirm our findings.

### Wake SW, a potential EEG marker for conversion to A+ in SCD

Wake SW amplitude was higher in A− individuals who later converted to A+ (A−_A+) compared to those who remained A− (A−_A−), but also compared to individuals already A+ at baseline (A+_A+). Although limited by its cross-sectional design, a previous study performed in the same cohort reported non-linear associations between amyloid load and theta and delta power, suggesting compensatory mechanisms at early stages of AD.^48^ Notably, delta power peaked at amyloid levels just below the threshold differentiating A− and A+, and to rise again at higher amyloid levels, consistent with the EEG slowing typically observed at later stages of the disease.^54^ Our models showed that wake SW amplitude alone is a good predictor of the conversion to A+, with notably high specificity. Many studies have explored EEG metrics to classify AD, MCI, and healthy controls, or to distinguish stable from progressive MCI.^55^ Despite the limited number of individuals converting to A+ during the follow-up (n=16), the present study stands out by targeting an earlier disease stage (i.e., SCD) and predicting the conversion to A+ using novel EEG metrics (i.e., wake SW features). Our findings suggest that wake SW is a promising prognostic marker of amyloid conversion in cognitively unimpaired individuals with memory complaints. Early identification of at-risk individuals is essential for targeted interventions and risk/benefit assessment, as highlighted by the aducanumab controversy.^56^ Future longitudinal studies in larger cohorts of A− to A+ converters are needed to confirm the prognostic potential of wake SW.

### Wake SW amplitude at rest reflects cognitive functioning

We found that higher amplitude of wake SW at rest correlated with poorer cognitive performance. This was observed across several cognitive domains, including memory, attention, and executive functions, as assessed by composite scores derived from neuropsychological tests. Previous literature in rodents and young adults showed that wake SW features during behavioral tasks can predict errors and slowed cognitive processing at the trial level,^29,32–34^ likely due to the neuronal silencing associated with wake SW.^29,34^ Notably, in these studies, the location of wake SW was related to the brain regions engaged by the task and predicted the type of behavioral error.^32,33^ Our findings suggest that resting-state wake SW can also provide information about cognition, although the partial overlap of our clusters across cognitive domains indicates limited specificity compared to studies linking task-related wake SW and behavior.

### Potential mechanisms modulating wake SW in SCD

We linked wake SW amplitude to 1) poorer cognition, 2) A+N+ status, and to 3) increased risk of conversion to A+ in elderly with SCD. Whether these wake SW arise from the same physiological mechanisms as those identified in rodents and young adults (where they have been interpreted as local intrusions of sleep^36^) or instead reflect age- or pathology-related processes, remains an open question. In our SCD population, increasing amyloid burden could generate inflammation,^57^ modify neuronal excitability,^58^ and disrupt cortico-cortical connectivity,^59^ which in turn might modulate neuronal synchronization and wake SW. Interestingly, inflammatory processes following brain injury have been shown to promote wake SW in cortical areas surrounding the lesion.^60^ These postlesional sleep-like SW may be partly triggered by the shared molecular regulators of inflammation and sleep.^61^ However, it remains unclear whether such wake SW are a ‘passive’ sign of brain suffering, or a compensatory mechanism with neuroprotective and restorative functions,^62,63^ two interpretations that are not necessarily mutually exclusive. Notably, while sleep SW are thought to play several important roles, such as in brain waste clearance,^26,64^ the functions of wake SW are still unclear. A recent preprint reported that optogenetic induction of sleep-like ON/OFF periods during wakefulness (i.e., the bistable activity pattern underlying SW) produces similar effects on molecular markers of cortical excitatory synaptic strength and memory consolidation as natural sleep.^65^ Additionally, recent findings indicate that sleep deprivation enhances pulsatile CSF flow in young adults,^66,67^ accompanied by increased delta power, impaired attention and reduced behavioral performance.^67^ If confirmed with direct measures of brain clearance, this would mean that under certain circumstances, some cortical cleaning mechanisms could be partially re-activated during wakefulness, but at a cost: more SW and less attention. In this scenario, the transient rise in wake SW amplitude could be a compensatory mechanism to amyloid accumulation that becomes overwhelmed at higher amyloid levels.^48^ Further studies are needed to directly quantify potential metabolite clearance associated with wake SW and the cognitive consequences on the AD continuum.

### Wake SW and sex-differences

In our exploratory analyses, we found that men had higher wake SW amplitude (in delta and theta bands), as well as higher posterior SW density (in theta band only) compared to women. To our knowledge, sex-differences in wake SW have not been previously examined. Our findings should therefore be interpreted with caution, particularly given the gender imbalance in our dataset. If replicated, the observed group differences could be considered in the context of the potential role of wake SW in brain clearance. From this perspective, the wake SW pattern observed in men may compensate for their faster age-related decline in N3 stage and sleep SW.^68,69^ Interestingly, CSF clearance-related processes measured at rest show sex-differences after age 55, with women displaying sharper declines that coincide with menopausal hormonal change.^70^ Greater resilience to AD pathological processes have also been reported in men,^71^ while women typically exhibit an initial advantage in cognitive abilities, before experiencing a faster decline.^72^ This aligns with our observation of elevated wake SW amplitude in men. If future studies confirm our finding in older adults and show no sex-differences in wake SW before menopause, this would strengthen the hypothesis that wake SW serve a compensatory function, or arise as a by-product of compensatory processes.

## Conclusion

Our study highlights the potential of wake SW as a meaningful prognostic biomarker in the context of AD. We demonstrate that such EEG metrics can help identify at-risk individuals during the earliest disease stages, prior to cognitive decline and even when the amyloid burden is low. Importantly, wake SW can be measured during a simple and short resting-state EEG session, an inexpensive, non-invasive, and accessible technique. Further research could extend these investigations to later stages of AD to better understand the full trajectory of wake SW dysregulations.

## Data Availability

The data that support the findings of this study are available from the corresponding author, upon reasonable request.

## Acknowledgements

We thank all the staff at the Paris Brain Institute (ICM) and at the institute for memory and Alzheimer’s disease (IM2A) who supported the INSIGHT-preAD project. We acknowledge all the INSIGHT-preAD volunteers and the members of the INSIGHT-preAD research group (listed in Supplementary file). We also thank the Fondation Recherche Alzheimer for their financial support. Finally, we are grateful to Arthur Le Coz for the insightful discussions on wake SW, and to Nathalie George for her valuable help on EEG preprocessing.

## Consent Statement

The ethics committee of the Pitié-Salpêtrière University Hospital approved the study protocol. Written informed consent according to the Declaration of Helsinki was provided by all participants.

