## Supplementary files for "Sleep-like slow waves during resting-state: a promising EEG biomarker of amyloid and neurodegeneration in preclinical Alzheimer’s disease"

**List of the members of the INSIGHT-preAD study group:**

Audrain C, Auffret A, Bakardjian H, Baldacci F, Batrancourt B, Benakki I, Benali H, Bertin H, Bertrand A, Bombois S, Boukadida L, Cacciamani F, Causse V, Cavedo E, Cherif Touil S, Chiesa PA, Chupin M, Colliot O, Dalla Barba G, Depaulis M, Dos Santos A, Dubois B, Dubois M, Epelbaum S, Fontaine B, Francisque H, Gagliardi G, Genin A, Genthon R, Glasman P, Gombert F, Habert MO, Hampel H, Hewa H, Houot M, Jungalee N, Kas A, Kilani La Corte V, Le Roy F, Lehericy S, Letondor C, Levy M, Lista S, Lowrey M, Ly J, Makiese O, Mangin JF, Masetti I, Mendes A, Metzinger C, Michon A, Mochel F, Nait Arab R, Nyasse F, Perrin C, Poirier F, Poisson C, Potier MC, Ratovohery S, Revillon M, Rojkova K, Santos-Andrade K, Schindler R, Servera MC, Seux L, Simon V, Skovronsky D, Thiebaut M, Uspenskaya O, Villain N, Vlaincu M, Younsi N.

### Supplementary figures

**Figure S1: Topographies of wake SW features.**

**Figure S2: Effect of A and N status on the distribution of wake SW amplitude.**

**Figure S3: Changes in wake SW features between the pre- and post-task resting-state sessions for each A/N group.**

**Figure S4: Effect of ongoing amyloid dynamic on the distribution of wake SW amplitude.**

**Figure S5: Evolution of resting-state wake SW features from baseline to 2-year later.**

**Figure S6: Effect on ongoing amyloid dynamic on resting-state wake SW features quantified after the transition to A+ (at year-2 follow-up).**

### Supplementary tables

**Supplementary Table 1. Characteristics of each A/N group at the year-2 follow up.**

**Supplementary Table 2. Characteristics of each A_A group at baseline.**

**Supplementary Table 3. Prediction of individuals who will convert from A− to A+ during the 2-year follow-up period, after controlling for sex.**

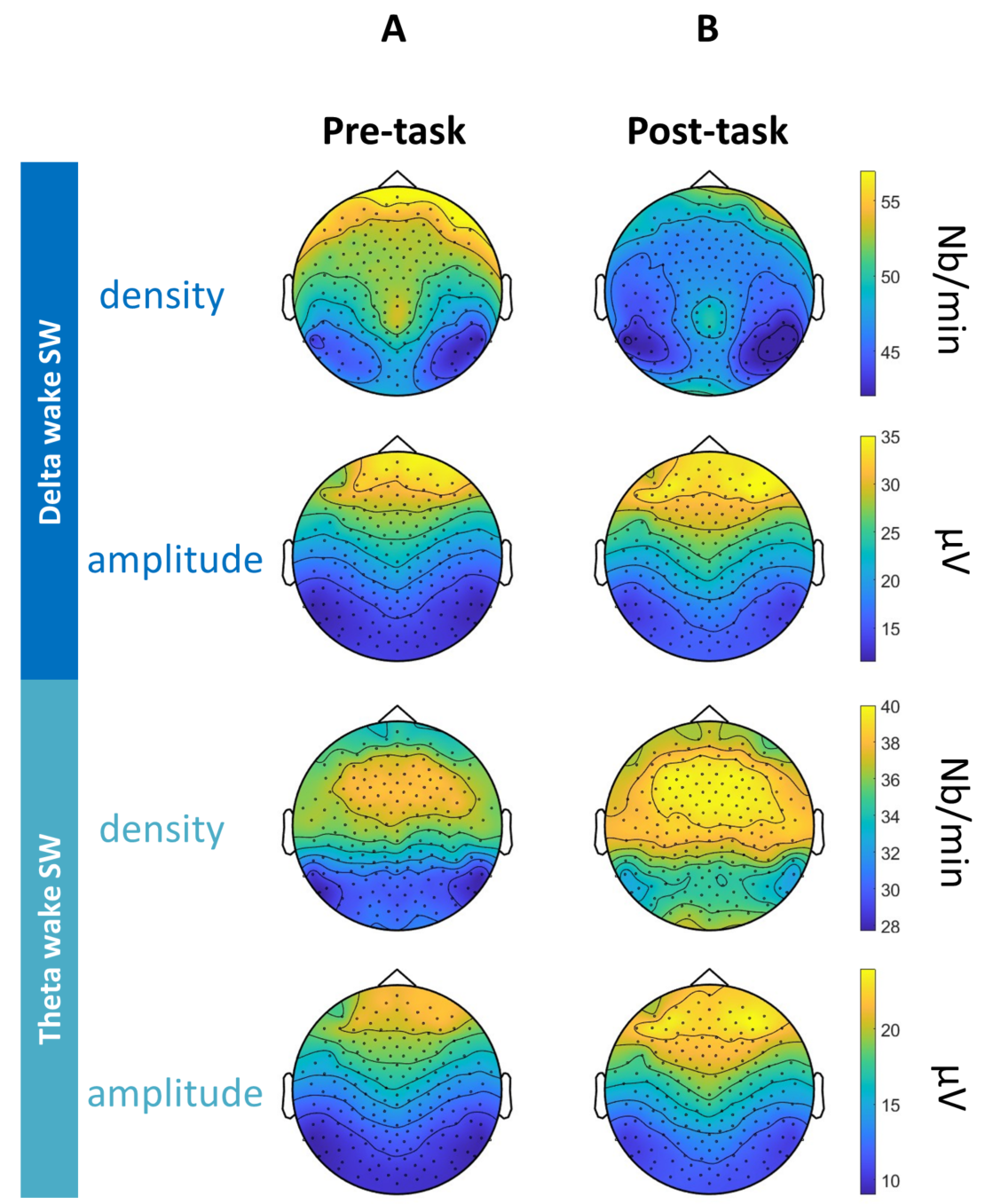

**Figure S1: Topographies of wake SW features.** *Topographies representing mean wake SW features (density and amplitude) during the pre-task and post-task resting-states (average across all participants).*

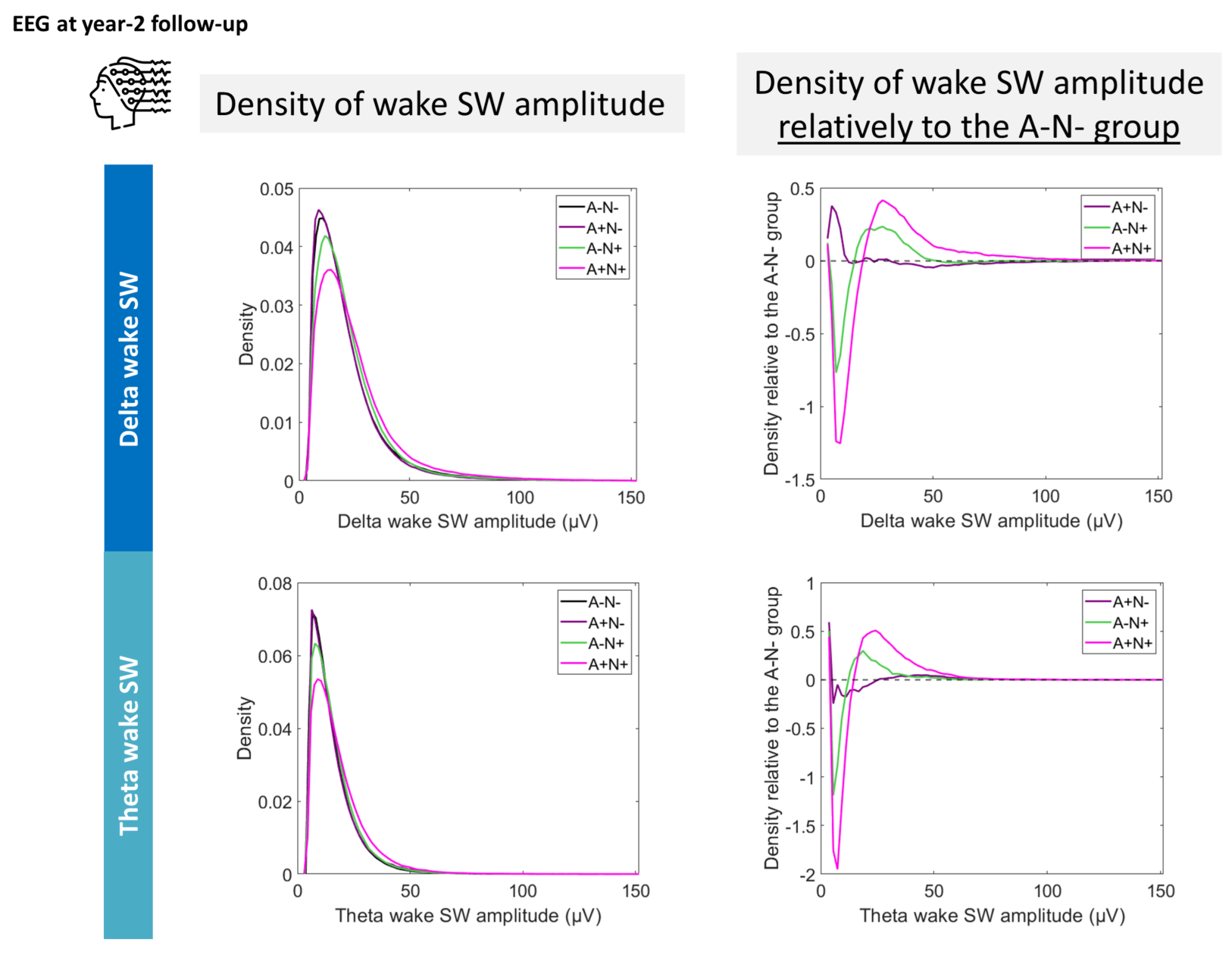

**Figure S2: Effect of A and N status on the distribution of wake SW amplitude.** *Density distribution of the wake SW amplitude during resting-state recordings (data from all channels have been pooled). Left panels show absolute density, while right panels show density relative to the A−N− group (i.e., density of the group of interest minus density of the A−N− group). Data at the year-2 follow-up were used as group distributions were more balanced compared to baseline (****Table 1****). Abbreviations: A (Amyloid), N (Neurodegeneration), SW (Slow Waves).*

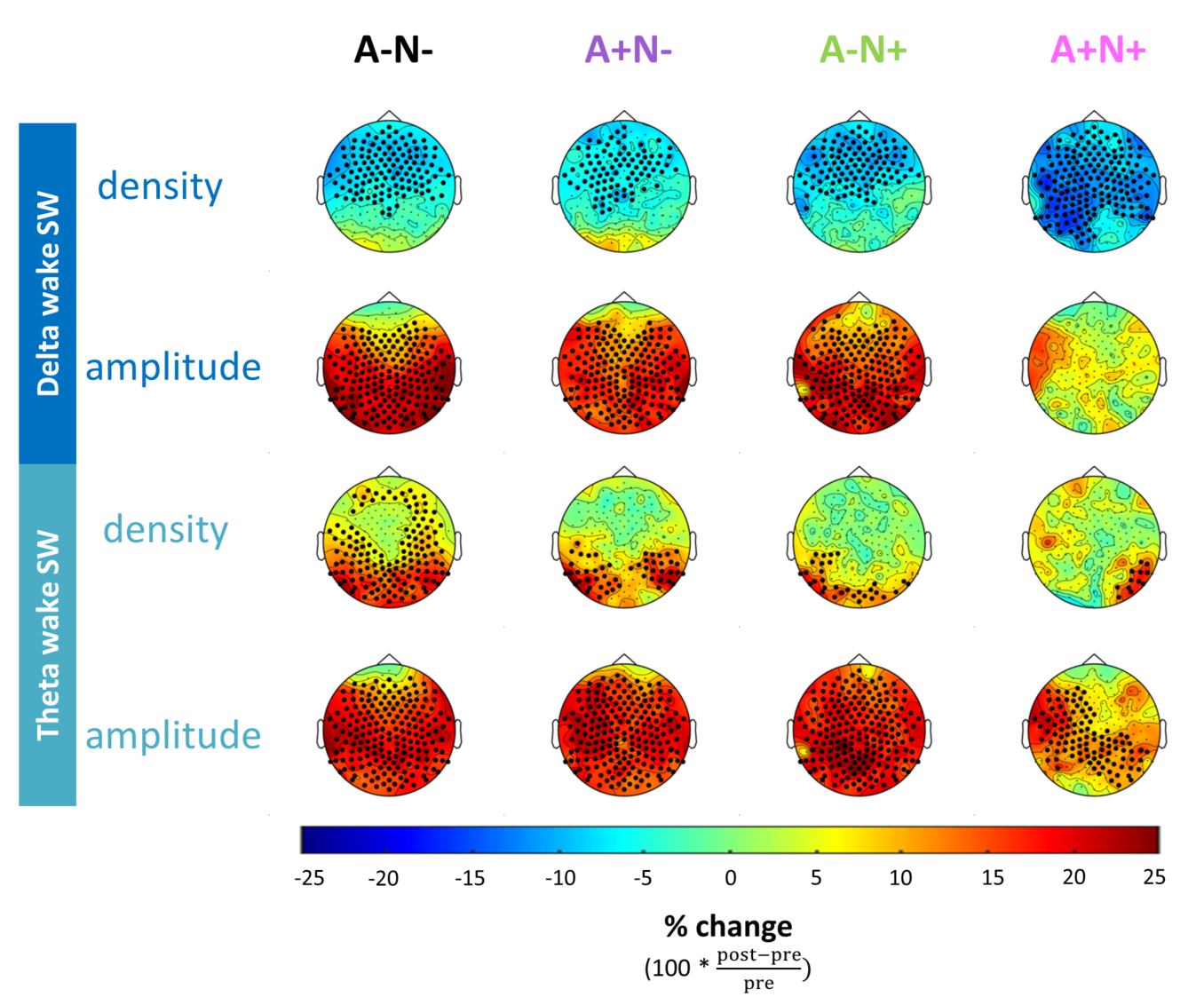

**Figure S3: Changes in wake SW features between the pre- and post-task resting-state sessions for each A/N group.** *Topographies representing the proportional change in mean wake SW metrics (density and amplitude) between the two resting-state recordings, for each A/N group. Data at the year-2 follow-up were used as group distributions were more balanced compared to baseline (Table 1). Pre- and post-task EEG data were available for 98 A–N–, 40 A+N–, 42 A–N+, and 29 A+N+ participants. Positive values indicate an increase in the post-task relative to the pre-task resting-state, while negative values denote a decrease. Black dots (•) represent significant clusters of electrodes using a cluster-based permutation approach after controlling for age, sex and education level using linear mixed models. Abbreviation: A (Amyloid), N (Neurodegeneration), SW (Slow Waves).*

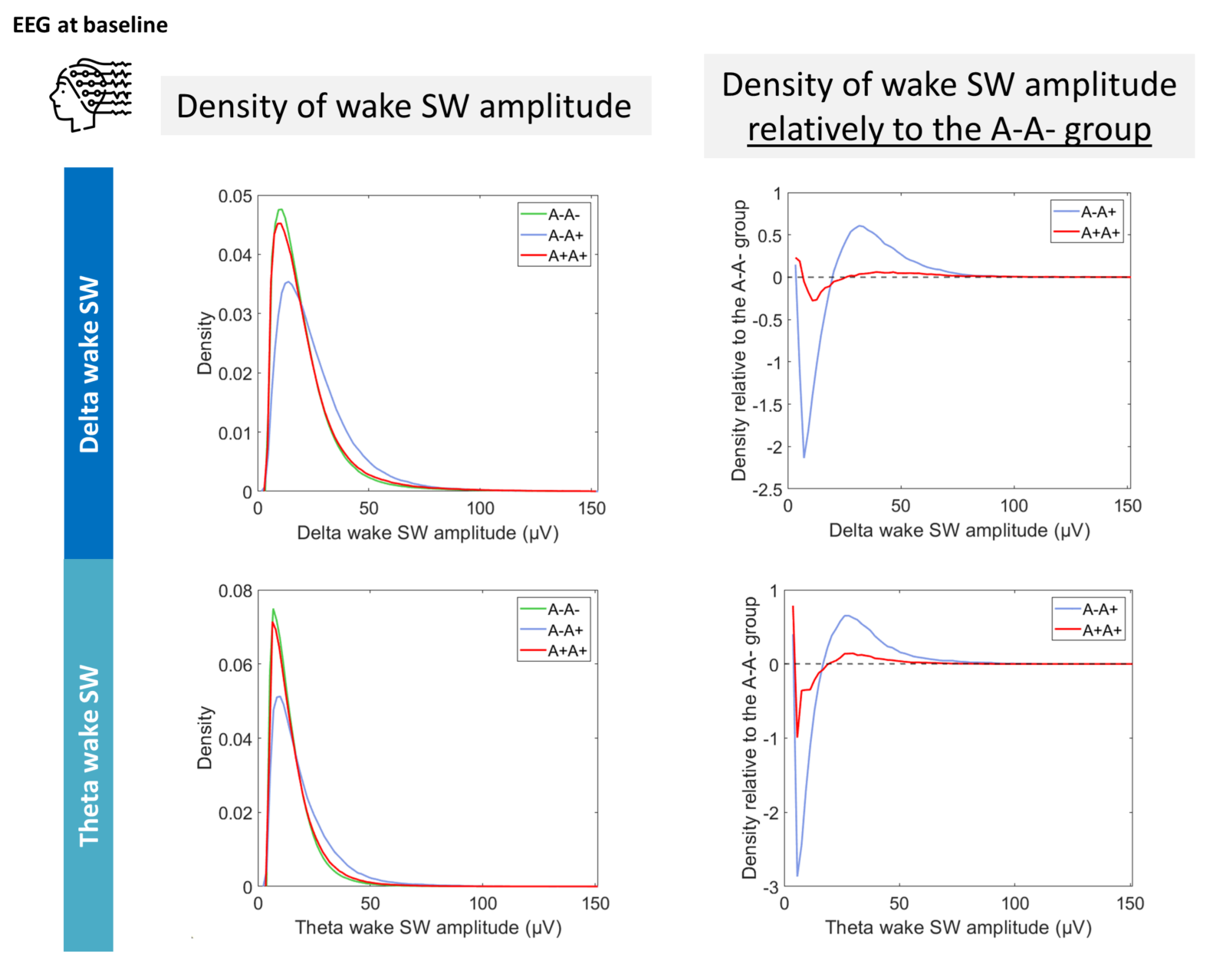

**Figure S4: Effect of ongoing amyloid dynamic on the distribution of wake SW amplitude.** *Density distribution of the wake SW amplitude during resting-state recordings (data from all channels have been pooled). Left panels show absolute density, while right panels show density relative to the A−_A− group (i.e., density of the group of interest minus density of the A−_A− group). Abbreviations: A (Amyloid), SW (Slow Waves).*

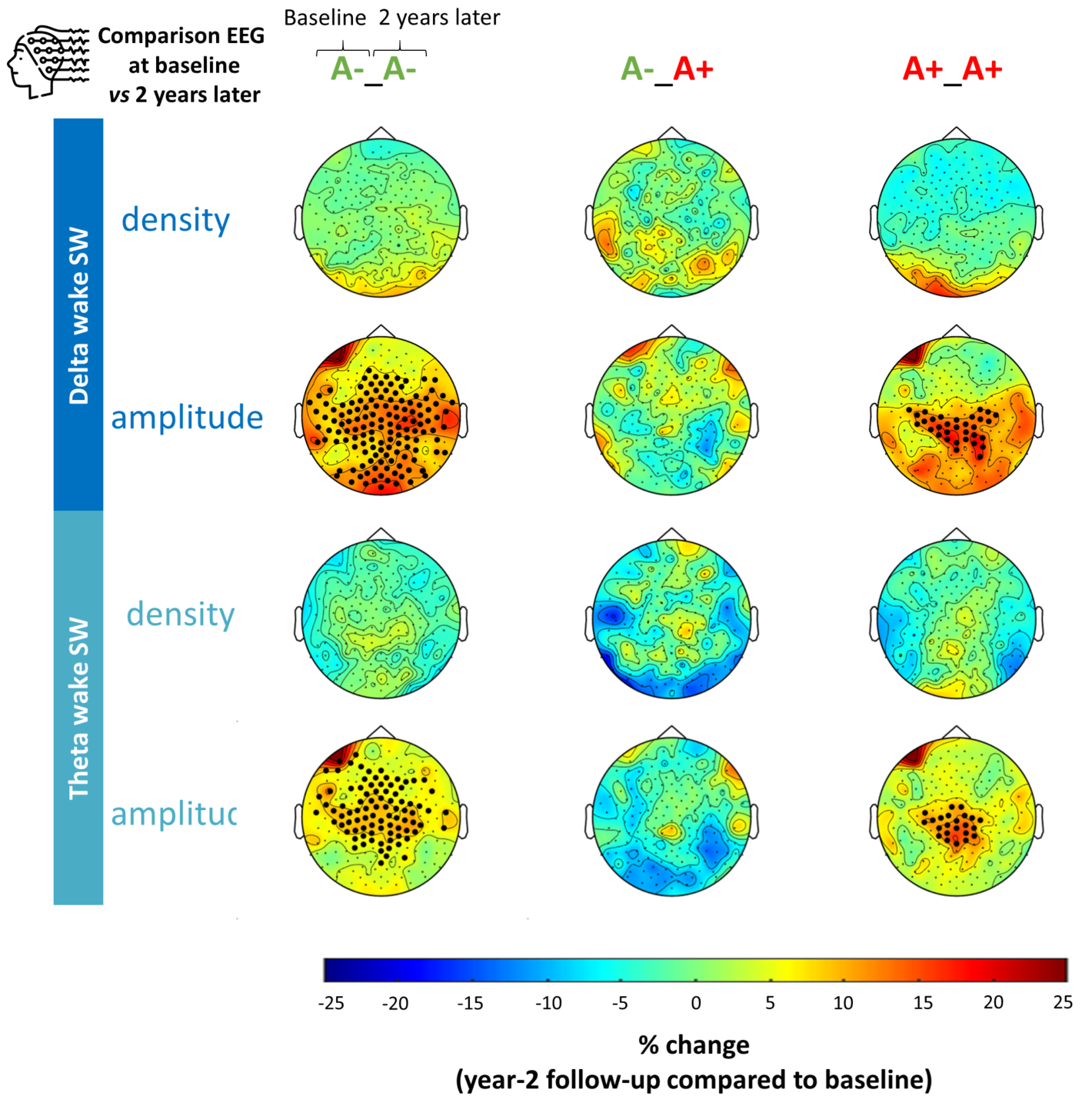

**Figure S5: Evolution of resting-state wake SW features from baseline to 2-year later.** *Topographies representing the proportional change in mean resting-state wake SW metrics (density and amplitude) over the 2-year follow-up for each group, stratified by the evolution of their A status (A−_A− group, A−_A+ group, and A+_A+ group) (e.g.,* $100* \frac{\left( mean SW amplitude at year-2 follow-up \right)-\left( mean SW amplitude at baseline \right)}{mean SW amplitude at baseline}$*within the A−_A− group). Positive values indicate an increase at year-2 follow-up relative to baseline, while negative values denote a decrease. Black dots (•) represent significant clusters of electrodes using a cluster-based permutation approach after controlling for age, sex and education level using linear models. Abbreviations: A (Amyloid), SW (Slow Waves).*

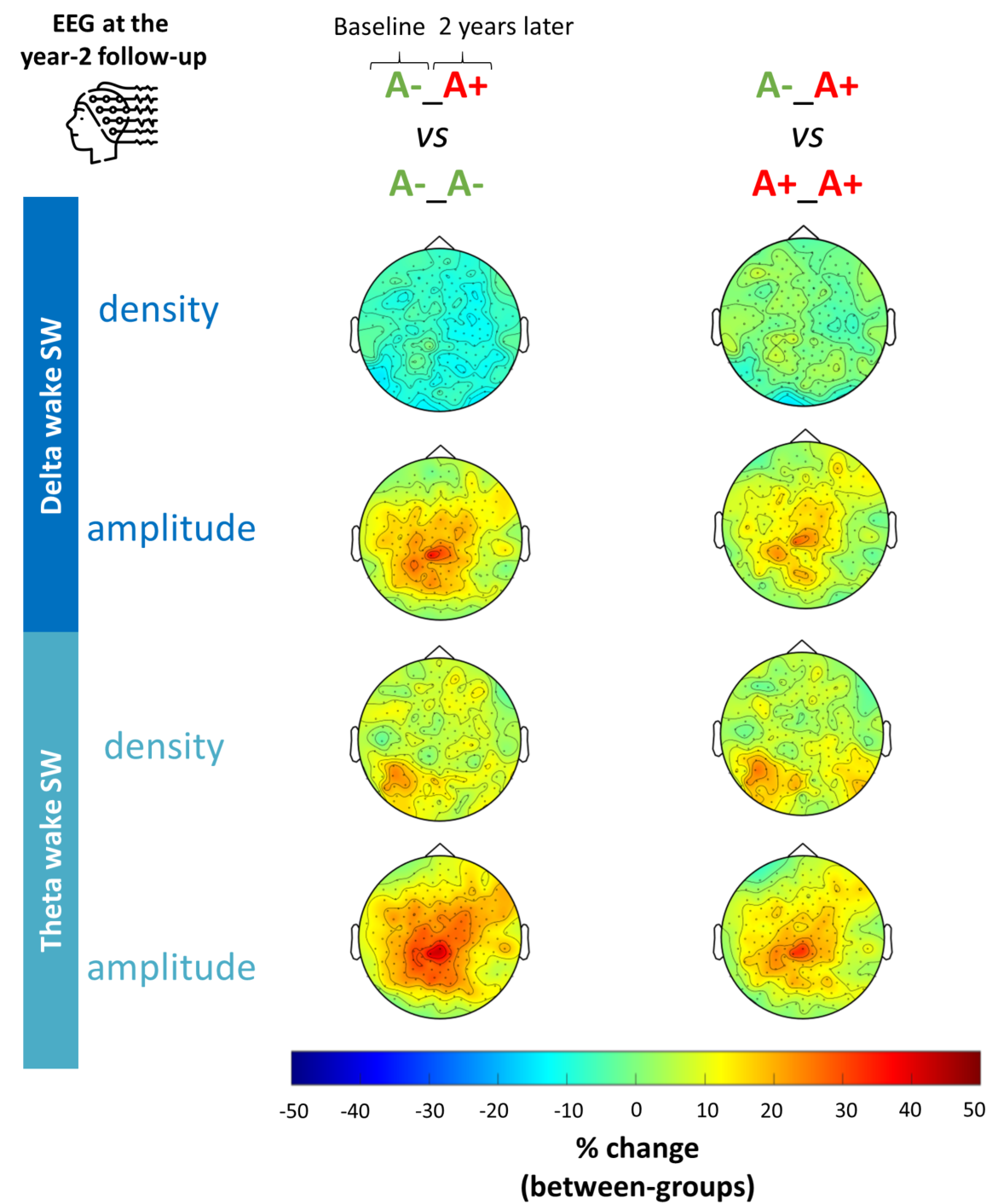

**Figure S6: Effect on ongoing amyloid dynamic on resting-state wake SW features quantified after the transition to A+ (at year-2 follow-up).**

*Topographies representing the proportional change in mean resting-state wake SW metrics (density and amplitude, quantified at year-2 follow-up) in individuals who converted to amyloid positivity over the 2-year follow-up (A−_A+ group) compared to those who remained either amyloid negative (A−_A− group) or positive (A+_A+ group) (e.g.,* $100* \frac{\left( mean SW amplitude in A-\_A+ group \right)-\left( mean SW amplitude in A-\_A-group \right)}{mean SW amplitude in A-\_A-group}).$*Positive values indicate higher density (or amplitude) in the A−_A+ group, while negative values denote lower density (or amplitude). Black dots (•) represent significant clusters of electrodes using a cluster-based permutation approach after controlling for age, sex and education level using linear models. Abbreviations: A (Amyloid), SW (Slow Waves).*

**Supplementary Table 1. Characteristics of each A/N group at the year-2 follow up.**

|  | **A−N−**  **(a)** | **A+N−**  **(b)** | **A−N+**  **(c)** | **A+N+**  **(d)** | **p-value** |
| --- | --- | --- | --- | --- | --- |
| **Demographics** |  |  |  |  |  |
| Number of individuals | 109 | 45 | 43 | 37 | / |
| Age (years) | 77.8 ± 3.3 | 77.5± 2.8 | 79.2± 3.7 | 79.2 ± 3.6 | **0.022** |
| Sex ratio (% of women) | 67.9^d^ | 66.7 | 53.5 | 40.5 | **0.016** |
| Education (% of individuals with high education) | 75.2 | 60.0 | 69.8 | 75.7 | 0.258 |
| ApoE4 genotype (% of ApoE4 carriers) | 14.7^b,d^ | 31.1^a,c^ | 4.7^b,d^ | 43.2^a,c^ | **< 0.001** |
| **Neuroimaging data** |  |  |  |  |  |
| Mean SUVr ^18^F-florbetapir | 0.62 ± 0.04^b,d^ | 0.82 ± 0.10 ^a,c,d^ | 0.61 ± 0.03 ^b,d^ | 0.90 ± 0.15 ^a,b,c^ | **< 0.001** |
| Mean SUVr ^18^F-FDG | 2.16 ± 0.13 ^c,d^ | 2.12 ± 0.10 ^c,d^ | 1.88 ± 0.08 ^a,b^ | 1.83 ± 0.11 ^a,b^ | **< 0.001** |
| **Cognition** |  |  |  |  |  |
| MMSE (total score, /30) | 28.96 ± 1.21 | 28.93 ± 1.21 | 28.58 ± 1.45 | 28.46 ± 1.43 | 0.114 |
| FCSRT (immediate total score, /48) | 46.84 ± 1.69 | 46.82 ± 1.47 | 46.40 ± 1.80 | 45.97 ± 2.85 | 0.078 |
| FAB (total score, /18) | 16.96 ± 1.32 ^d^ | 16.58 ± 1.32 ^d^ | 16.77 ± 1.48 ^d^ | 15.57 ± 2.29 ^a,b,c^ | **< 0.001** |

*Data are presented as mean ± standard deviation for continuous metrics. Characteristics of A/N groups are indicated at the year-2 follow-up as group distributions were more balanced compared to baseline (****Table 1****). Thirteen participants could not be classified into one of the four groups due to missing ^18^F-florbetapir-PET or ^18^FDG-PET data. Group comparisons were performed using ANOVA for continuous variables and with a Chi^2^ test of independence for categorical variables. Superscript letters (a,b,c,d) denote groups that differ significantly from one another following post-hoc pairwise comparisons with Tukey's HSD correction (for continuous variables) and Benjamini–Hochberg correction (for categorical variables). Abbreviations: A (Amyloid), FAB (Frontal Assessment Battery at Bedside), FCSRT (Free and Cued Selective Reminding Test), FDG (FluoroDeoxyGlucose), MMSE (Mini-Mental State Examination), N (Neurodegeneration), SUVr (Standardized Uptake Value ratio).*

**Supplementary Table 2. Characteristics of each A_A group at baseline.**

|  | **A−_A−**  **(a)** | **A−_A+**  **(b)** | **A+_A+**  **(c)** | **p-value** |
| --- | --- | --- | --- | --- |
| **Demographics** |  |  |  |  |
| Number of individuals | 157 | 16 | 63 | / |
| Age (years) | 76.3 ± 3.5 | 75.7 ± 3.8 | 76.5 ± 3.2 | 0.669 |
| Sex ratio (% of women) | 63.1^b^ | 31.3^a,c^ | 65.1^b^ | **0.035** |
| Education (% of individuals with high education) | 72.0 | 75.0 | 60.3 | 0.208 |
| ApoE4 genotype (% of ApoE4 carriers) | 12.7^c^ | 25.0 | 38.1^a^ | **< 0.001** |
| **Neuroimaging data** |  |  |  |  |
| Mean SUVr ^18^F-florbetapir | 0.60 ± 0.03^b,c^ | 0.65 ± 0.02 ^a,c^ | 0.86 ± 0.13 ^a,b^ | **< 0.001** |
| Mean SUVr ^18^F-FDG | 2.13 ± 0.22 | 2.07 ± 0.20 | 2.10 ± 0.20 | 0.329 |
| **Cognition** |  |  |  |  |
| MMSE (total score, /30) | 28.7 ± 0.98 | 28.9 ± 0.89 | 28.4 ± 0.93 | 0.113 |
| FCSRT (immediate total score, /48) | 46.1 ± 2.01 | 45.8 ± 2.14 | 46.1 ± 2.04 | 0.799 |
| FAB (total score, /18) | 16.6 ± 1.57 | 16.9 ± 0.89 | 16.1 ± 1.76 | 0.092 |

*Data are presented as mean ± standard deviation for continuous metrics. Characteristics of each group are indicated at baseline. Seven participants could not be classified into one of the three groups due to missing ^18^F-florbetapir-PET data at baseline or follow-up. Group comparisons were performed using ANOVA for continuous variables and with a Chi^2^ test of independence for categorical variables. Superscript letters (a,b,c,d) denote groups that differ significantly from one another following post-hoc pairwise comparisons with Tukey's HSD correction (for continuous variables) and Benjamini–Hochberg correction (for categorical variables). Abbreviations: A (Amyloid), FAB (Frontal Assessment Battery at Bedside), FCSRT (Free and Cued Selective Reminding Test), FDG (FluoroDeoxyGlucose), MMSE (Mini-Mental State Examination), N (Neurodegeneration), SUVr (Standardized Uptake Value ratio).*

**Supplementary Table 3. Prediction of individuals who will convert from A− to A+ during the 2-year follow-up period, after controlling for sex.**

|  | **Sensitivity**  **(TP rate)** | **Specificity**  **(TN rate)** | **Balanced accuracy** |
| --- | --- | --- | --- |
| Delta wake SW amplitude + Sex | 0.55 [0.47-0.62] | 0.87 [0.85-0.89] | 0.71 [0.67-0.73] |
| Theta wake SW amplitude alone + Sex | 0.57 [0.50-0.63] | 0.89 [0.86-0.90] | 0.73 [0.69-0.76] |
| Baseline ^18^F-florbetapir PET SUVr + Sex | 0.83 [0.73-0.90] | 0.73 [0.68-0.77] | 0.78 [0.72-0.81] |
| Baseline ^18^F-florbetapir PET SUVr + delta wake SW amplitude + Sex | 0.81 [0.68-0.90] | 0.78 [0.73-0.82] | 0.79 [0.73-0.83] |
| Baseline ^18^F-florbetapir PET SUVr + theta wake SW amplitude + Sex | 0.80 [0.67-0.93] | 0.79 [0.76-0.83] | 0.79 [0.73-0.86] |
| Baseline ^18^F-florbetapir PET SUVr + delta wake SW amplitude +  theta wake SW amplitude + Sex | 0.74 [0.60-0.87] | 0.78 [0.72-0.85] | 0.76 [0.70-0.81] |

*Conversion from A− to A+ status was modeled using logistic regression with baseline predictors (^18^F-florbetapir PET SUVR and/or wake SW amplitude in delta/theta bands), controlling for sex, using 100 iterations of 5-fold cross-validation. Mean values and 95% confidence intervals (in brackets) are reported for sensitivity, specificity and balanced accuracy (accounting for class imbalance). Abbreviations: TN (True Negative), TP (True Positive), SUVr (Standardized Uptake Value ratio).*
